# Day-to-Day Sleep Efficiency and Driving Behaviors in Older Adults with and without Cognitive Impairment

**DOI:** 10.1101/2025.02.11.25322072

**Authors:** Jun Ha Chang, Yunwen Huang, Ying Zhang, Su Chen, Daniel L. Murman, Vaishali Phatak, Matthew Rizzo

## Abstract

**INTRODUCTION:** Sleep disturbances are common in older adults, particularly those with cognitive impairment. This study examines how day-to-day sleep quality impacts real-world driving behaviors, offering insights into sleep as a functional biomarker of cognitive health.

**METHODS:** We monitored 149 community-dwelling older adults (90 cognitively impaired, 59 unimpaired) over 12 weeks. Sleep was measured via wrist-worn actigraphy and driving data via an in-vehicle sensor system. A zero-inflated Poisson regression model examined whether sleep efficiency was associated next-day driving likelihood and frequency, and whether these relationships varied by cognitive status.

**RESULTS:** Better sleep efficiency increased the likelihood of driving the following day more among cognitively impaired than unimpaired participants. Higher sleep efficiency was associated with increased driving frequency in both groups.

**DISCUSSION:** These findings underscore the importance of daily sleep variability as a potential digital biomarker for functional abilities in older adults, highlighting opportunities for early intervention to preserve mobility and independence.

## 1. BACKGROUND

Alzheimer’s disease and related dementias (ADRD) are leading causes of cognitive and functional decline in older adults. Emerging evidence highlights the critical role of sleep disturbances in exacerbating cognitive and functional impairments associated with ADRD. Reduced sleep efficiency and increased variability are linked to accelerated neurodegeneration and may serve as early indicators of disease progression.^1–4^ Characterizing and quantifying the effects of sleep disturbances on daily mobility and activities of daily living can offer valuable insights into the real-world signs and costs of ADRD.

Sleep disturbances affect up to 40% of individuals with ADRD and mild cognitive impairment (MCI).^4–6^ Poor sleep quality and sleep deprivation can impair memory, executive functioning, and emotional regulation, contributing to cognitive decline.^4,7^ Growing evidence highlights the impact of day-to-day variability in sleep on neurodegeneration and cognitive performance.^8,9^ Baril et al. (2024)^10^ found that greater variability in sleep patterns (e.g., sleep midpoint, sleep duration) was associated with abnormal AD biomarkers, including altered cerebrospinal fluid (CSF) ratios of p-*τ*181 to amyloid-*β*42, suggesting that sleep fluctuations may reflect underlying neurodegenerative processes. Balouch et al. (2022)^11^ found that increased night-to-night sleep variation predicted day-to-day changes in cognitive and behavioral functions, such as heightened daytime sleepiness, reduced alertness, memory errors, and behavioral problems in individuals with AD and MCI. These studies underscore the critical importance of understanding day-to-day real-world sleep variability as an indicator and a modifiable contributor to cognitive decline and reduced quality of life.^12^ The real-world functional consequences of this variability require quantification and further detail.

Automobile driving serves as a powerful lens for assessing real-world cognitive and behavioral function in individuals at risk for ADRD.^13–15^ As a complex, safety-critical, and mobility-enhancing activity, driving depends on a range of cognitive and motor processes, including attention, decision-making, executive function, and reaction time—all of which are highly sensitive to factors such as sleep quality, medications, and others. Older adults with MCI are particularly at risk for, as cognitive deficits combined with poor sleep can exacerbate driving impairments. This increased vulnerability may lead to self-imposed or externally recommended restrictions on driving, ultimately reducing mobility, independence, and quality of life.

This study examined the day-to-day relationships between sleep quality, measured via wrist-worn actigraphy, and naturalistic driving behaviors captured through an in-vehicle sensor system in older adults with and without cognitive impairments. We tested the hypothesis that poorer sleep quality, defined as lower sleep efficiency relative to an individual’s baseline, would be associated with reduced driving mobility the following day, particularly among those with cognitive impairment––as detailed in the Methods below.

## 2. METHODS

### 2.1 Participants

Community-dwelling older adults were enrolled in longitudinal studies of Alzheimer’s Disease (AD) and driving behaviors at the University of Nebraska Medical Center (UNMC) Mind and Brain Health Labs between March 2021 and January 2024. Potential participants completed pre-screening to assess if they met the eligibility criteria for inclusion into the study: 1) age between 65 and 90 years inclusive and living independently in Nebraska or nearby (e.g. Iowa); 2) held a valid driver’s license; 3) able to communicate in English; and 4) capable of informed consent.

Exclusion criteria were: 1) a pulmonary disease requiring chronic medication; 2) congestive heart failure; 3) a major psychiatric illness; 4) active vestibular disease; 5) substance abuse within the year prior to enrollment; 6) use of medications that could confound the study results; and 7) evidence of medical or neurologic conditions (e.g., stroke), as judged by a clinician, that could confound the study results. The study was approved by the UNMC Institutional Review Board (0522-20-FB), and all participants signed an informed consent.

### 2.2 Procedure

Figure 1 shows the flow chart of the study procedure. At induction, participants completed a demographic questionnaire and started to monitor their real-world sleep and driving behaviors via wrist-worn actigraphy and in-vehicle driving sensor devices on their vehicles, respectively. After around 2 weeks, participants visited the site with their study partner to assess cognition and daily functioning. After 12 weeks since the study induction, participants visited the site to return the actigraphy and uninstall the driving sensor devices.

**Figure 1.**
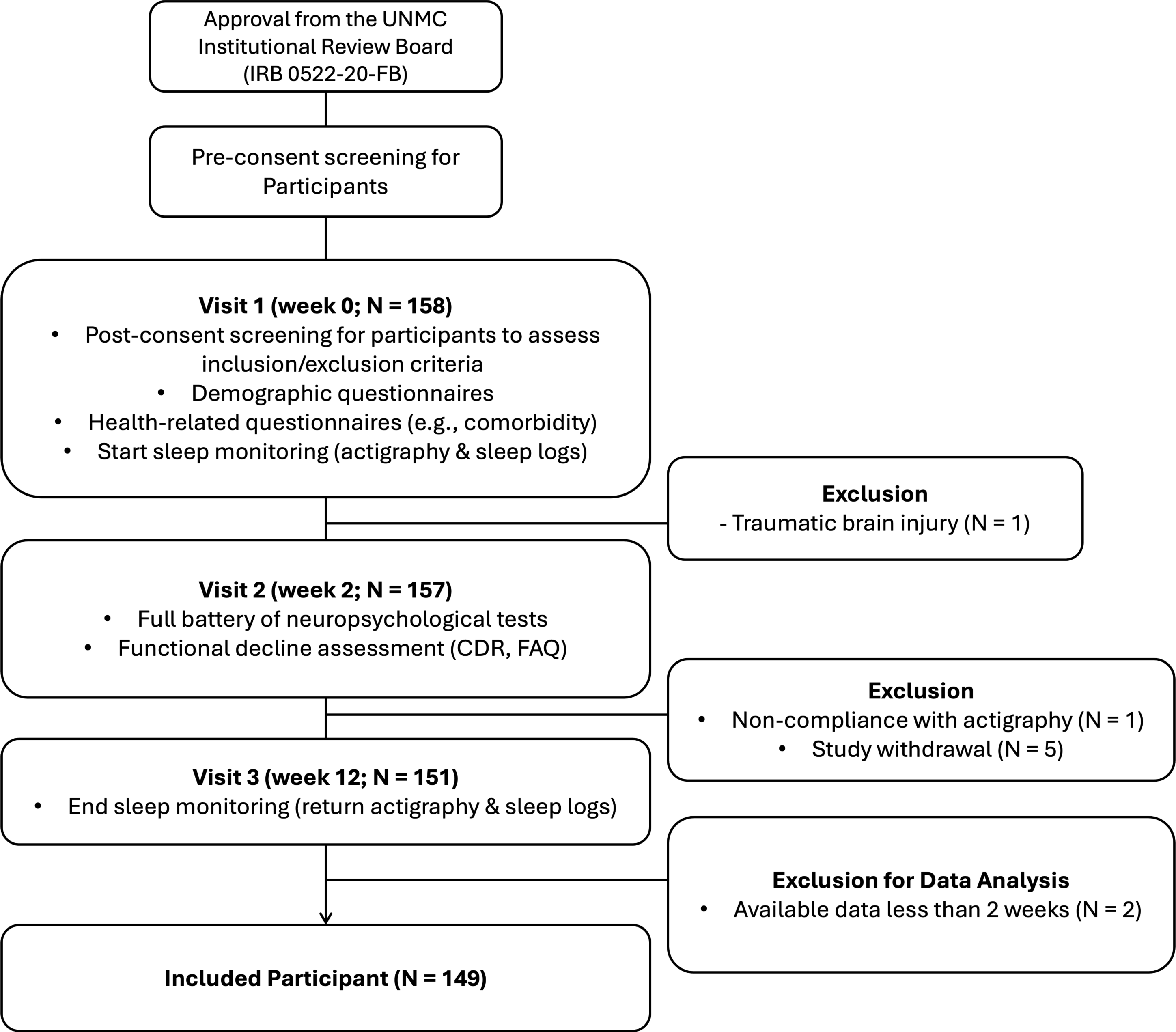
Flowchart illustrating participant recruitment, screening, and data collection processes. Participants were screened for eligibility, consented, and monitored for both sleep and driving behaviors over 12 weeks.

### 2.3 Cognitive Status

We followed the 2018 NIA-AA research criteria for syndromal staging of the cognitive continuum.^16^ Clinicians who specialized in dementia and AD reviewed neurological and neuropsychological assessments for cognition and daily functioning from NACC UDS version 3^17^ and reached a consensus on cognitive staging. To determine cognitive status, participants completed tests spanning five domains: (1) memory, (2) language, (3) visuospatial skills, (4) executive function, and (5) attention. All specific tests are presented in Supplementary Table 1. Raw scores were converted to age-, sex-, and education-adjusted z-score. These z-scores were used to define the presence of mild cognitive impairment (MCI) according to both Peterson/Winblad MCI criteria (at least one cognitive test impaired per domain, > 1.5 z-score below expectation)^18,19^ and Jak/Bondi MCI criteria (at least two cognitive tests per domain, > 1 z-score below expectation).^20^ Also, functional status was assessed using Functional Activities Questionnaires (FAQ)^21^ and Clinical Dementia Rating Scale (CDR® Dementia Staging Instrument)^22^. Consistent with diagnostic criteria, a global CDR score of ≧ 0.5^22^ and FAQ score > 9^23^ were required to determine significant impairment in cognition and instrumental activities of daily living (IADLs). Given both cognitive and functional decline, individuals classified as cognitively unimpaired (CU) showed no cognitive or functional decline, whereas those with cognitive deficits without substantial functional decline were categorized as MCI. When cognitive decline co-occurred with functional decline, participants were considered to have dementia. Due to relatively small sample size of dementia (N = 9), we created a group called cognitively impaired (CI) consisting of participants with MCI and dementia.

### 2.4 Sleep

Participants wore the ActiGraph GT9X tri-axial accelerometer (ActiGraph LLC, 30Hz sampling rate) on their non-dominant wrist for 12 consecutive weeks, except for water-related activities, concurrently recording the time to go to bed and time to out of bed on paper sleep logs. Study partners can assist, if needed, to ensure the device was worn and the sleep logs were consistently filled out. Sleep data from the actigraphy were downloaded two weeks post-consent and at the end of 12 weeks.

Sleep variables were derived by aligning log-based bedtimes with actigraphy data and scoring 1-minute epochs as sleep or wake using the Cole-Kripke algorithm that validated for older adults against polysomnography.^24^ Wear-time was assessed with the Choi algorithm, which detects non-wear epochs and is validated for older adults.^25^ We focused on two nocturnal sleep variables: total sleep time (TST), calculated by summing sleep epochs, and sleep efficiency (SE), calculated as the ratio of TST to the total spent in bed to estimate sleep duration and quality, respectively. Nights with device non-wear or abnormal sleep periods that defined by multiple sleep periods separated by gaps exceeding 1 hour in a single night, were excluded (7.5%) to avoid misrepresentation.

### 2.5 Naturalistic Driving

A custom-built vehicle sensor instrumentation system (Black Box) was installed at the study start in each participant’s vehicle to collect real-world driving data for 12 consecutive weeks concurrently with wearing the actigraphy. The system continuously and passively recorded video (forward roadway, cabin), global positioning system (GPS), accelerometer, and on-board diagnostic (engine throttle, rpm, and speed) data every second from on- to off-ignition. The system was unobstructively mounted on the forward windshield, next to the rearview mirror. Each participant was asked to drive as they typically would.

We excluded driving trips if the distance driven was 0 miles (on- and off-ignition recorded), and the maximum speed during the trip was less than 5 miles/hour (8.05 km/hour) as considering driving in parking lots or driveway (5.2%). Also, certain periods when participants reported that they could not drive or had no driving data available due to various reasons (e.g., vacations, COVID-19 quarantine, Black Box system malfunction) were also excluded (7.4%).

### 2.6 Data Cleaning and Preparation

Real-world sleep and driving data were combined on a day-to-day basis, ensuring that only days with both driving and sleep data were included in the analysis. Two participants with fewer than 2 weeks of valid data were excluded (6 and 7 nights, respectively). After applying these criteria, the final dataset included a total of 10,291 participant-days with an average of 69 days of data per participant, used for further analyses.

### 2.7 Statistical Analysis

Descriptive statistics for demographic, disease-specific measures, and actigraphy-measured sleep were assessed to determine baseline participant characteristics and compared the CU and CI groups using independent *t*-tests for continuous variables and Chi-square test for categorical variables.

#### 2.7.1 Primary analysis

We aimed to determine whether SE the night before affects driving behaviors and if this relationship varied by cognitive status. The **outcome variable** was the number of driving trips per day (count), with SE and cognitive group (CU and CI) as **primary predictors.**

To account for individual differences in sleep patterns, SE and TST were converted into z-scores based on each participant’s mean and standard deviation. This transformation allows values to be interpreted relative to a participant’s typical sleep pattern, with a z-score of 0 representing the mean, -2 indicating atypically poor sleep, and +2 indicating better-than-average sleep. Other continuous variables, such as age and education, were standardized at the group level to ensure consistency across variables measured on different scales.

Given the repeated measures, where multiple observations were recorded for each participant, we included a **random effect for participants** to account for within-subject correlation. In addition, the model was adjusted for the following **covariates**: age, gender, season (winter [November – March] vs. non-winter [April– October]) with a reference level of not-winter, and working status (working vs. not-working) with a reference level of not-working.

Upon reviewing the distribution of the outcome variable, we found it was right-skewed with many days having no driving trips. To account for this excess of zero values, we used a longitudinal zero-inflated Poisson (ZIP) regression model.^26,27^ This model addresses accounts for two distinct underlying the data: (1) a binary model accounting for the likelihood of no driving and (2) a Poisson model for the counts of driving trips on days. By explicitly modeling these two processes, ZIP regression more accurately captured the observed distributions of days with and without driving trips.

#### 2.7.2. Sensitivity Analysis

We conducted sensitivity analyses to evaluate whether TST moderates the relationship between SE and driving behaviors across cognitive groups. First, we compared models with and without TST using the Likelihood Ratio Test (LRT), Akaike Information Criterion (AIC), and Bayesian Information Criterion (BIC). Improved model fit with TST inclusion justified its addition as a predictor. Then, we replaced the two-way interaction term (SE x cognitive group) with a three-way interaction term (SE x cognitive group x TST) to assess whether TST moderates the relationship among driving behavior, SE and cognitive groups. If the three-way interaction was not significant, we retained TST as an independent predictor for preserving model parsimony.

## 3. RESULTS

### 3.1 Participant characteristics

Table 1 summarizes the baseline characteristics of participants, stratified by cognitive status. No significant differences were observed in age, education, employment status, season, or medication use for sleep disorders. However, the CU group included more females compared to the CI group (*p* = 0.018), while the CI group had a higher proportion of African American participants (*p* = 0.017). These differences are discussed further in the Discussion section.

**Table 1.**
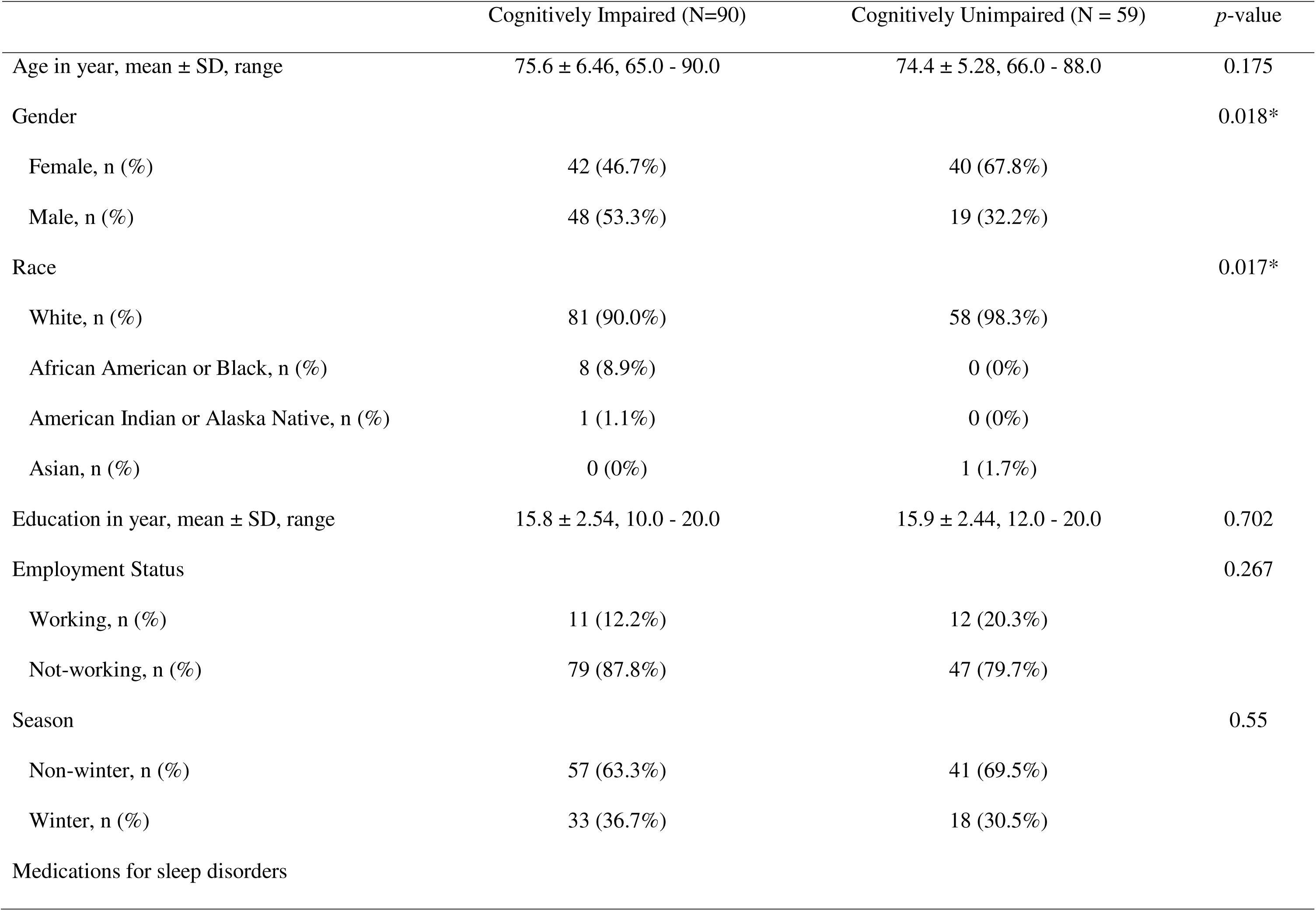
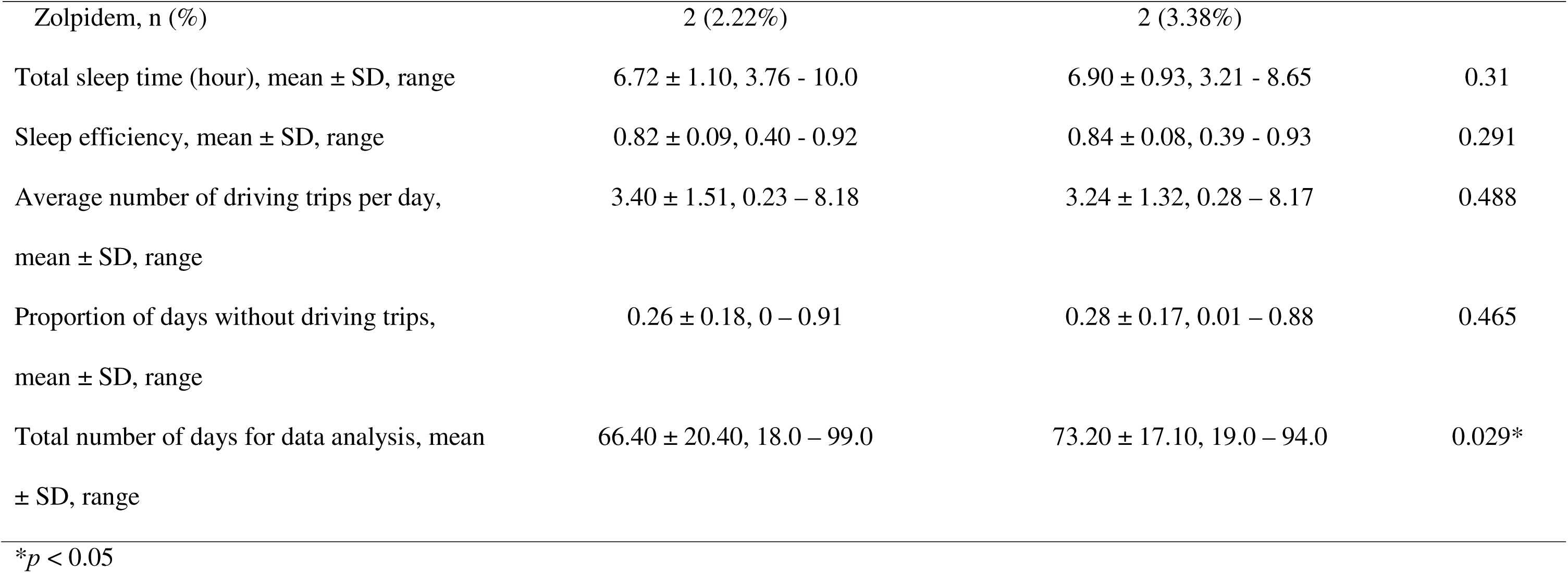
Participant Characteristics.

Sleep characteristics over two weeks were similar between groups, with average TST of 6.62 hours (CI) and 6.90 hours (CU), and sleep efficiency of 0.82 and 0.84, respectively. Valid data availability was approximately one week longer for the CU group (*p* = 0.029) due to higher actigraphy non-compliance in the CI group, such as forgetfulness in completing sleep diaries. Despite this, groups provided over two months of usable data.

### 3.2 Primary Results

#### 3.2.1 Poisson model

Table 2 (upper part) presents the results of the Poisson model, which evaluates the number of driving trips on days. We found that higher SE the night before was associated with a slight increase in the number of driving trips the next day (*Incidence Rate Ratio [IRR]* = 1.014, *95% Confidence Interval [CI]* = 1.001 – 1.026, *p* = 0.034, see Figure 2). In contrast, longer TST was associated with a decrease in the number of driving trips (*IRR* = 0.969, *95% CI* = 0.957 – 0.981, *p* < 0.001).

**Figure 2.**
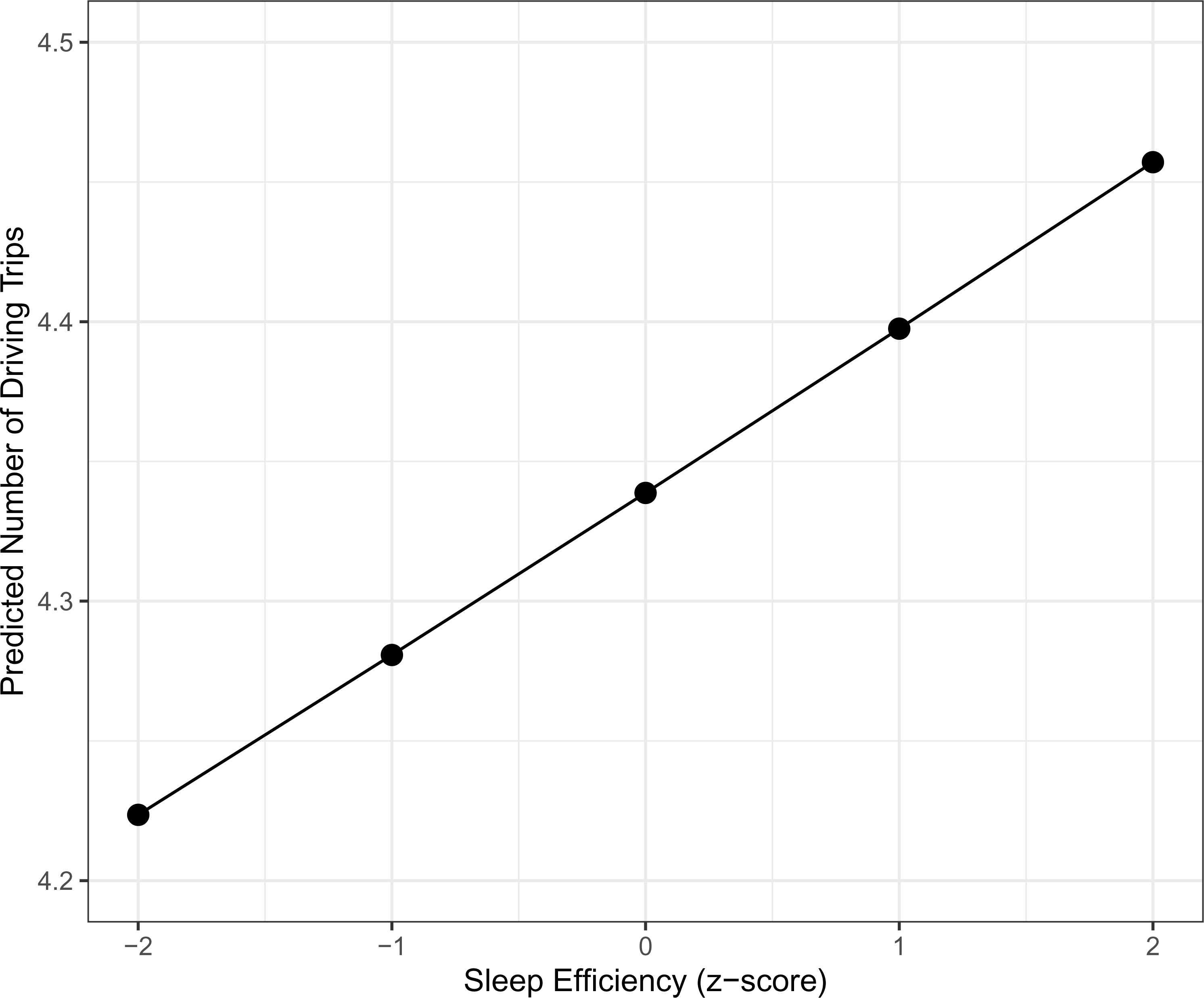
Predicted number of driving trips based on sleep efficiency. The x-axis represents sleep efficiency z-scores, with positive values indicating better sleep efficiency and negative values indicating worse sleep efficiency relative to the individual’s mean. The y-axis represents the predicted number of driving trips on the next day, derived from a Poisson model. Predictions are based on a reference individual with average sleep efficiency (z = 0), average age (z = 0), male, during the winter season, and employed. These reference values are used to facilitate interpretation and do not represent specific individuals in the dataset.

**Table 2.** Results of Zero-Inflated Poisson Model.

| Predictors | Incidence Rate Ratio (95% CI) | *p*-value |
| --- | --- | --- |
| Poisson Model |  |  |
| Cognitive Group (CI) | 1.022 (0.934 - 1.119) | 0.633 |
| Sleep Efficiency | 1.014 (1.001 - 1.026) | 0.034* |
| Total Sleep Time | 0.969 (0.957 - 0.981) | < 0.001*** |
| Age | 0.941 (0.901 - 0.981) | 0.005** |
| Gender (Male) | 1.025 (0.939 - 1.120) | 0.577 |
| Season (Winter) | 0.969 (0.933 - 1.007) | 0.113 |
| Working Status (Work) | 1.060 (0.942 - 1.193) | 0.333 |
| Zero-Inflated (Binary) Model |  |  |
| Predictors | Odds Ratio (95% CI) | *p*-value |
| Cognitive Group (CI) x Sleep Efficiency | 0.877 (0.791 - 0.973) | 0.013* |
| Cognitive Group (CI) | 0.800 (0.556 - 1.150) | 0.228 |
| Sleep Efficiency | 0.960 (0.884 - 1.042) | 0.329 |
| Total Sleep Time | 1.241 (1.169 - 1.317) | < 0.001*** |
| Age | 1.075 (0.903 - 1.280) | 0.415 |
| Gender (Male) | 0.955 (0.668 - 1.365) | 0.8 |
| Season (Winter) | 1.288 (1.088 - 1.525) | 0.003** |
| Working Status (Work) | 0.572 (0.351 - 0.931) | 0.024* |

\* $p < 0.05$ , \*\* $p < 0.01$ , \*\*\* $p < 0.001$

The cognitive status groups were not significantly associated with the number of driving trips (*IRR* = 1.022, *95% CI* = 0.934 – 1.119, *p* = 0.633), neither was the interaction between the cognitive group and SE, which was therefore excluded from the final model. Age, however, was a significant factor, with older participants demonstrating fewer driving trips (*IRR* = 0.941, *95% CI* = 0.901 – 0.982, *p* = 0.005). There were no significant associations found for gender, employment status, and season.

#### 3.2.2. Zero-inflated (binary) model

Table 2 (lower part) presents the results of the zero-inflated model, which evaluates the likelihood of no driving on a given day. First and most importantly, there was a significant interaction effect between the cognitive group and SE (*Odds Ratio [OR]* = 0.877, *95% CI* = 0.791 – 0.973, *p* = 0.013), indicating that the relationship between SE and the likelihood of no driving differed based on cognitive status.

As illustrated in Figure 3, the CU group showed relatively stable likelihood of no driving across different z-scores of SE, with only a slight increase in the likelihood of no driving as SE decreased. In contrast, the CI group showed a more pronounced effect that better SE significantly reduced the likelihood of no driving. For example, at a z-score of -2 (indicating poor SE), the predicted probability of no driving was substantially higher for the CI group compared to the CU group.

**Figure 3.**
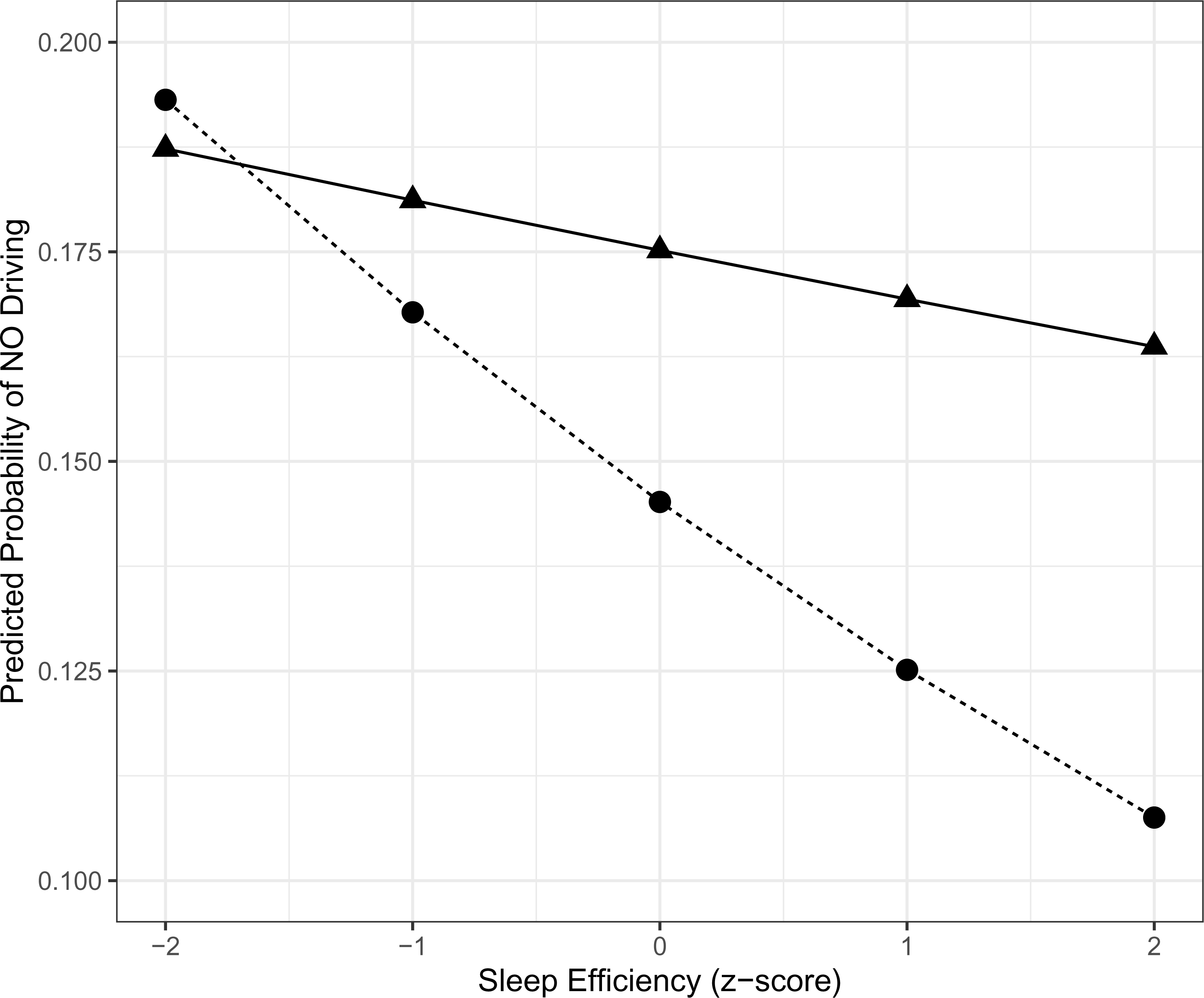
Predicted probability of no driving based on sleep efficiency. The x-axis **represents** sleep efficiency z-scores, with positive values indicating better sleep efficiency and negative values indicating worse sleep efficiency relative to the individual’s mean. The y-axis represents the predicted probability of no driving on the next day, where lower values correspond to a higher likelihood of driving. Results are stratified by cognitive status: CU (Cognitively Unimpaired) and CI (Cognitively Impaired). Predictions are based on a reference individual with average sleep efficiency (z = 0), average age (z = 0), male, during the winter season, and employed. These reference values are used to facilitate interpretation and do not represent specific individuals in the dataset.

Additionally, increased TST was associated with a higher likelihood of no driving (*OR* = 1.241, *95% CI* = 1.169 – 1.317, *p* < 0.001). The winter season was also significantly associated with an increased likelihood of no driving (*OR* = 1.288, *95% CI* = 1.088 – 1.525, *p* = 0.003). Participants who were employed were less likely to have no driving (*OR* = 0.572, *95% CI* = 0.351 – 0.931, *p* = 0.024). None of other variables significantly affects the likelihood of no driving the following day.

### 3.3 Sensitivity Analysis

Model comparisons demonstrated that the inclusion of TST significantly improved model performance compared to the model without TST (χ*^2^*=80.275, *p* < 0.001). This improvement was also reflected by lower AIC (with TST: 43,746 vs. without TST: 43,822) and BIC (with TST: 43,883 vs. without TST: 43,945), indicating that TST provides meaningful explanatory power for driving behaviors and should be included in the final model.

However, the three-way interaction (TST × SE × Cognitive Status) was not significant in the zero-inflated component (*OR* = 0.985, *95% CI* = 0.906 – 1.071, *p* = 0.728). This result suggests that TST does not moderate the relationship among driving, SE, and the cognitive group. Instead, TST appears to exert an independent, direct effect on driving behaviors.

## 4. DISCUSSION

This study aimed to examine how sleep quality and quantity relate to day-to-day driving behaviors among older adults with and without cognitive impairment. To achieve this, we combined objective sleep measures with naturalistic driving data collected over three months. By integrating these measures, we evaluated whether sleep characteristics independently affect driving patterns and how cognitive status may modify these associations.

Two critical findings from our study highlight the interplay between cognitive status, sleep, and driving behaviors in older adults. First, while higher sleep efficiency (SE) was associated with an increase in the number of driving trips on days participants chose to drive, longer total sleep time (TST) correlated with a higher likelihood of no driving at all. This suggests that sleep quality, rather than sleep duration alone, may be important for sustaining the cognitive alertness and functional capacity required for daily driving.^28–31^ Previous studies have similarly found that individuals with better sleep quality often demonstrate better daytime functioning and decision-making abilities,^32^ whereas excessive or prolonged sleep can sometimes be linked to underlying health issues including dementia^33–35^ and reduced engagement in daily activities.^36^ Our results align with literature that emphasizes the importance of sleep quality over mere quantity to help maintain functional outcomes and independence in older adults.

Second, the effect of SE on driving likelihood was more pronounced among participants with cognitive impairment (CI) compared to those without (CU). While the CU group was relatively resilient to fluctuations in sleep quality, the CI group were more sensitive: poorer SE increased their likelihood of refraining from driving the next day, whereas better SE increased their driving likelihood. These findings align with emerging evidence that even early-stage cognitive impairment can amplify the risk factors on daily functioning like driving.^37–39^ Although previous studies have focused primarily on visual or attentional deficits as key factors in driving cessation or restrictions among older adults with neurodegenerative conditions^40–42^, our findings extended this line of research by highlighting sleep quality as another modifiable factor affecting real-world driving decisions. In doing so, this study provides additional empirical evidence to a growing body of literature that underscores the multifactorial nature of driving behaviors in aging populations and suggests that improving sleep efficiency may help preserve mobility and independence among older adults facing dementia.

These results also suggest that improved sleep may temporarily mitigate cognitive deficits, enhancing decision-making and motor coordination. Slow-wave and REM sleep, which are crucial for memory and emotional regulation and are often disrupted in individuals with MCI or early-stage dementia,^43,44^ may explain this effect. The CI group may also demonstrate better self-regulation by avoiding driving on poor-sleep days and utilizing better-rested days, unlike the CU group, whose cognitive resilience supports stable driving patterns regardless of sleep quality.

We did not observe a significant interaction between cognitive status and SE regarding the number of driving trips taken once participants intend to drive. Instead, the interaction primarily affected the likelihood of no driving at all, underscoring the idea that sleep quality can affect the threshold for driving decision, particularly for those with cognitive impairment, but may not differentially affect the intensity of driving activity once underway.

Our results also suggest that longer sleep durations were associated with a reduced driving frequency and a higher likelihood of no driving. This finding may initially appear counterintuitive. However, extended sleep durations may reflect compensatory behavior for poor-quality rest or underlying health issues such as depression,^45^ and might simply reduce available daytime hours for activities like driving. Our sensitivity analyses support the latter idea. Although TST improved the overall power of our models, it did not alter how day-to-day SE interacts with cognitive impairment to affect the likelihood or frequency of driving, suggesting TST and SE have independent effects on driving activity. TST may shape broader daily activity patterns, whereas SE has a more direct effect on day-to-day driving decisions, especially in older adults with cognitive impairment.

Our findings highlight the potential of continuous, real-time monitoring of sleep and driving behaviors as a tool for early detection of disease progression. Real-world data on sleep patterns and functional activities could help identify early signs of cognitive decline, paving the way for timely interventions.

By integrating objective sleep data with real-world functional outcomes, this study provides new insights into how daily sleep variations influence functional decisions in older adults. The observed impact of sleep variability on cognitive and functional outcomes supports the potential of continuous monitoring to inform future disease management strategies. Coupling these real-world cognitive and functional measures with biomarker assessments could help harmonize the International Working Group’s (IWG) clinical-biological definition of AD with the Alzheimer’s Association’s biomarker-focused approach, balancing symptom emphasis with early detection and intervention.^46,47^

### 4.1 Limitations and future directions

Several limitations should be acknowledged. First, the CI group included individuals with both MCI and mild dementia, potentially introducing variability in functional impairments. Larger samples in future studies could help examine subgroup-specific dynamics. Second, demographic imbalances between the CI and CU groups, such as higher representation of males and African Americans in the CI group, may have influenced results, as these groups are often associated with poorer sleep quality and greater sleep fragmentation,^48,49^ potentially affecting driving behaviors. Third, the participants were drawn from a specific geographic and demographic context, limiting generalizability to populations with different cultural, socioeconomic, or healthcare factors. Future research across more diverse samples is needed to improve external validity. Lastly, while participants reported being the primary drivers, some driving data may reflect trips made by others (e.g., spouse). Future studies could address this by using in-cabin video or computer vision algorithms to confirm driver identity.

Despite these challenges, this study is among the first to link real-world driving behaviors to sleep quality, providing a foundation for future research. Integrating data from diverse sources—such as sleep, physical activity, driving behaviors, and electronic health records—could support predictive systems for cognitive decline, enabling targeted and personalized interventions to preserve mobility, independence, and quality of life in older adults.

## Consent Statement

All participants provided and signed an informed consent.

## Data Availability

All data produced in the present study are not available due to participants' privacy.

## Acknowledgements

We appreciate and would like to acknowledge to the individuals who contributed significantly to the data collection and processing for this study. Special thanks go to Kellee Halliburton, Karla Lynch, Melissa Hatch, and the rest of the Mind & Brain Health Labs who played a crucial role in administering participant visits and collecting and annotating of real-world data. The NACC database is funded by NIA/NIH Grant U24 AG072122. NACC data are contributed by the NIA-funded ADRCs: P30 AG062429 (PI James Brewer, MD, PhD), P30 AG066468 (PI Oscar Lopez, MD), P30 AG062421 (PI Bradley Hyman, MD, PhD), P30 AG066509 (PI Thomas Grabowski, MD), P30 AG066514 (PI Mary Sano, PhD), P30 AG066530 (PI Helena Chui, MD), P30 AG066507 (PI Marilyn Albert, PhD), P30 AG066444 (PI John Morris, MD), P30 AG066518 (PI Jeffrey Kaye, MD), P30 AG066512 (PI Thomas Wisniewski, MD), P30 AG066462 (PI Scott Small, MD), P30 AG072979 (PI David Wolk, MD), P30 AG072972 (PI Charles DeCarli, MD), P30 AG072976 (PI Andrew Saykin, PsyD), P30 AG072975 (PI David Bennett, MD), P30 AG072978 (PI Neil Kowall, MD), P30 AG072977 (PI Robert Vassar, PhD), P30 AG066519 (PI Frank LaFerla, PhD), P30 AG062677 (PI Ronald Petersen, MD, PhD), P30 AG079280 (PI Eric Reiman, MD), P30 AG062422 (PI Gil Rabinovici, MD), P30 AG066511 (PI Allan Levey, MD, PhD), P30 AG072946 (PI Linda Van Eldik, PhD), P30 AG062715 (PI Sanjay Asthana, MD, FRCP), P30 AG072973 (PI Russell Swerdlow, MD), P30 AG066506 (PI Todd Golde, MD, PhD), P30 AG066508 (PI Stephen Strittmatter, MD, PhD), P30 AG066515 (PI Victor Henderson, MD, MS), P30 AG072947 (PI Suzanne Craft, PhD), P30 AG072931 (PI Henry Paulson, MD, PhD), P30 AG066546 (PI Sudha Seshadri, MD), P20 AG068024 (PI Erik Roberson, MD, PhD), P20 AG068053 (PI Justin Miller, PhD), P20 AG068077 (PI Gary Rosenberg, MD), P20 AG068082 (PI Angela Jefferson, PhD), P30 AG072958 (PI Heather Whitson, MD), P30 AG072959 (PI James Leverenz, MD).

## Figure Legends

**Figure 3.** Two groups are depicted: CU (Cognitively Unimpaired), represented by a solid line with filled triangles, and CI (Cognitively Impaired), represented by a dashed line with filled circles.

